# Identification and genomic characterisation of BA.3.2: a highly divergent BA.3-related SARS-CoV-2 lineage from southern Africa

**DOI:** 10.64898/2025.12.19.25342658

**Authors:** Graeme Dor, Dikeledi Kekana, Ryan Hisner, Darren P. Martin, Bette Korber, Timo Ernst, Avram Levy, David Speers, Stuart Turville, Vitali Sintchenko, Kerri Basile, Geraldine Sullivan, Rebecca Rockett, Jen Kok, Federico Gueli, Richard Lessels, Cheryl Baxter, Nicole Wolter, Anne von Gottberg, Houriiyah Tegally, Tulio de Oliveira

**Affiliations:** Centre for Epidemic Response and Innovation (CERI), School for Data Science and Computational Thinking, Stellenbosch University, Stellenbosch, South Africa; Centre for Respiratory Diseases and Meningitis, National Institute for Communicable Diseases (NICD), a division of the National Health Laboratory Service (NHLS), Johannesburg, South Africa; Division of Computational Biology, Department of Integrative Biomedical Sciences, Institute of Infectious Diseases and Molecular Medicine, University of Cape Town, Cape Town, South Africa; Los Alamos National Laboratory, Los Alamos, NM, USA; New Mexico Consortium, Los Alamos, NM, USA; PathWest Laboratory Medicine WA, Perth, Western Australia, Australia; School of Biomedical Sciences, University of Western Australia, Perth, Western Australia, Australia; School of Medicine, University of Western Australia, Perth, Western Australia, Australia; The Kirby Institute, University of New South Wales, Sydney, New South Wales, Australia; Centre for Infectious Diseases and Microbiology Laboratory Services, NSW Health Pathology-Institute of Clinical Pathology and Medical Research; Centre for Infectious Diseases and Microbiology-Public Health; University of Sydney; Westmead Hospital, Westmead, New South Wales, Australia; Independent Researcher, Como, Italy; KwaZulu-Natal Research Innovation and Sequencing Platform (KRISP), Nelson R. Mandela School of Medicine, University of KwaZulu-Natal, Durban, South Africa; School of Pathology, Faculty of Health Sciences, University of the Witwatersrand, Johannesburg, South Africa

## Abstract

In November 2024, a highly divergent BA.3-related SARS-CoV-2 lineage, designated BA.3.2, was detected in South Africa, marking the first appearance of a BA.3-derived lineage in over two years. Phylogenetic reconstruction places BA.3.2 on an extended branch descending from ancestral BA.3, with no intermediate genomes detected, consistent with a prolonged period of unsampled or isolated evolution. Molecular clock analyses indicate accelerated divergence characteristic of a saltation event, while phylogeographic and demographic analyses point to a southern African origin followed by multiple independent exportations and evidence of ongoing global transmission. Relative to ancestral BA.3, BA.3.2 harbours 39 amino acid substitutions in the spike glycoprotein, two N-terminal domain deletions (Δ136–147 and Δ243–244), a four-residue insertion (ins214:ASDT), and a large deletion spanning ORF7a, ORF7b, and ORF8. The co-occurrence of D405N and R408S implies epistasis between these sites, while reversions R493Q and H505Y likely enhance ACE2 binding and antibody escape. Extensive remodeling across the spike, including loss of the C15–C136 disulfide bond and substitutions in the SD1 and SD2 domains, may influence spike stability, cleavage, and fusogenicity. The emergence and continued circulation of BA.3.2 underscores the ongoing potential for highly divergent SARS-CoV-2 variants to arise and spread globally. Despite its limited prevalence, the persistence of BA.3.2 alongside dominant lineages, together with evidence of more recent expansion, indicates that this lineage retains the potential to become of epidemiological concern under favourable conditions.

## Introduction

The initial emergence of the SARS-CoV-2 Omicron Variant of Concern (VOC) in late 2021 marked a fundamental break in SARS-CoV-2 evolution. Three related but distinct Omicron lineages, BA.1, BA.2, and BA.3, appeared at approximately the same time. All possessed over 30 new spike mutations relative to their nearest ancestor, which had ceased to circulate more than one year prior^1^. BA.1 was the first Omicron variant to spread internationally and in many countries caused a rapid wave of infections that dwarfed all previous COVID-19 waves. The initial BA.1 wave was shortly followed by a BA.2 wave, which went on to spawn a multitude of sublineages including those that are presently dominating globally^2^.

BA.3 was the least fit of the three initial Omicron variants, with by far the weakest ACE2 affinity, even lower than that of the Wuhan reference spike, which was extremely unusual in the VOC era^3^. Only about 760 BA.3 sequences were ever recorded^4,5^, over half of which were collected in Poland. All of the BA.3 spike mutations were also found in either BA.1 or BA.2. The BA.3 spike NTD region (S:1-305) was identical to the BA.1 NTD with the exception of the BA.1 214:EPE insertion, while all residues downstream of the RBD (S:529-1273) were identical to BA.2. The BA.3 RBD, as well as the remainder of its genome, had an approximately even mixture of BA.1 and BA.2 mutations^1^. By May 2022, BA.3 had disappeared from circulation.

The reappearance of BA.3 in November 2024 was therefore entirely unexpected, even in the context of substantially reduced global genomic surveillance. First detected in South Africa, BA.3.2 represents yet another Omicron BA lineage with an origin in southern Africa, the same region where BA.1, BA.2 and BA.3 were first identified and where later divergent lineages such as BA.4, BA.5 and BA.2.86 also likely originated^6^. Beyond southern Africa, multiple detections of BA.3.2 have been reported in different parts of Europe, Australia, and the United States. The duration and diversity of these detections suggest that BA.3.2 has achieved a greater scale and persistence of spread than BA.2.87.1, the last highly divergent lineage to arise in southern Africa prior to the emergence of BA.3.2^7–10^.

Here, we report on the early detection and genomic analysis of BA.3.2. Besides phylogenetically examining the origins of BA.3.2 and inferring when and where the BA.3.2.1 and BA.3.2.2 sublineages likely arose, we examine whether the mutations that differentiate BA.3.2 from its nearest relatives are suggestive of it having originated within the context of a long-term infection.

## Results and Discussion

### Re-emergence of a highly divergent BA.3 descendant: BA.3.2

BA.3.2 was first detected in South Africa in November 2024, representing the first reappearance of a BA.3-related lineage in more than two years (Figure 1a). BA.3.2 lineage viruses have now been sampled across multiple countries, most frequently in South Africa, Australia, and Germany (Figure 1b), but also in the Netherlands, the USA, Mozambique, Denmark, Ireland, Scotland, and Slovenia. In South Africa, BA.3.2 lineages have persisted but have not expanded to become highly prevalent during 2025. Rather, the lineages most commonly sampled in the country have been descendants of the XFG lineage that has been common in the Americas and Europe throughout 2025 (Figure S2a).

**Figure 1:**
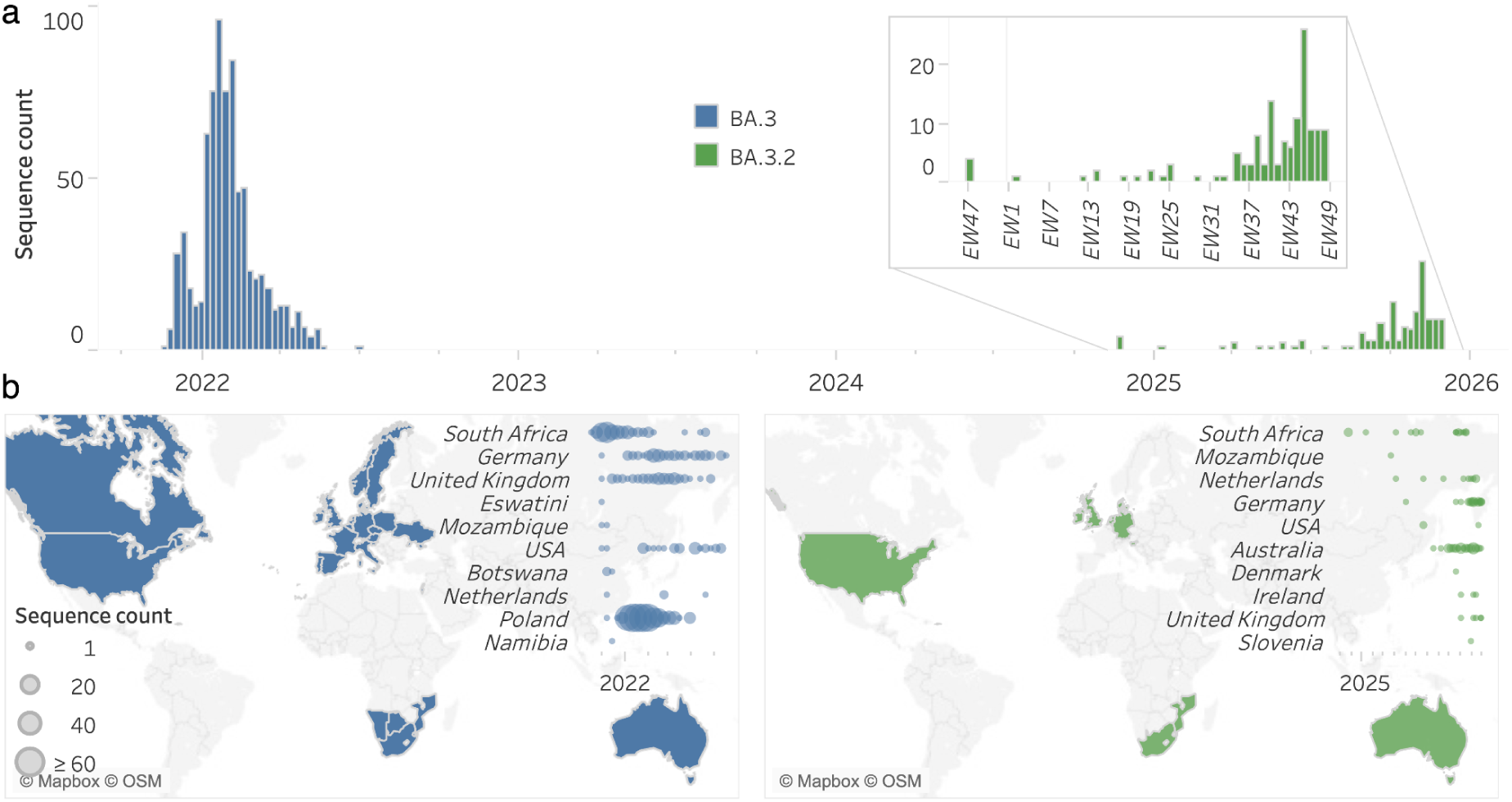
Weekly spatio-temporal genomic distribution of BA.3 lineages globally. **a** Time-series distribution of BA.3 sequences by epidemiological week (EW), stratified by Pango lineage (BA.3 shown in blue, BA.3.2 shown in green). The inset provides an expanded view beginning from the first detection of BA.3.2 in November 2024. **b** Geographic distribution of BA.3 (blue) and BA.3.2 (green) detections, with dot plots reflecting weekly sequence counts for each country. Countries are ordered by the date of first detection, and limited to the ten earliest-reporting countries.

Initial detections of BA.3.2 outside of South Africa only began occurring after April 2025 (2 April in the Netherlands; Figure 1b) with sequences being sporadically sampled in southern Africa and in various European countries over the next four months (14 sequences in total representing <1% of those sampled in these regions). Then, in July 2025, BA.3.2 sequences were detected in the Australian state of Western Australia (Figure S2d) marking the start of the biggest spike in BA.3.2 detections that we have currently seen. BA.3.2 is presently more prevalent in Australia than any other sampled region globally. Thus, this highly divergent lineage has achieved slow but persistent circulation for over a year, has spread across the world and has demonstrated its potential for accelerated spread in some regions, but has failed to rapidly increase in prevalence throughout the world in the way that previous saltation-variants such as BA.1, BA.2, and JN.1 did.

We phylogenetically reconstructed the evolutionary origins of BA.3.2 and its parent lineage, BA.3, by examining a representative subset of global SARS-CoV-2 genomes. Within this phylogenetic tree (Figure 2a) BA.3.2 is situated on a long, isolated branch descending from ancestral BA.3 sequences, with no intermediate genomes sampled during the intervening period: a pattern indicating that the progenitor BA.3.2 lineage underwent a prolonged phase of unsampled evolution between approximately December 2021 and November 2024.

**Figure 2.**
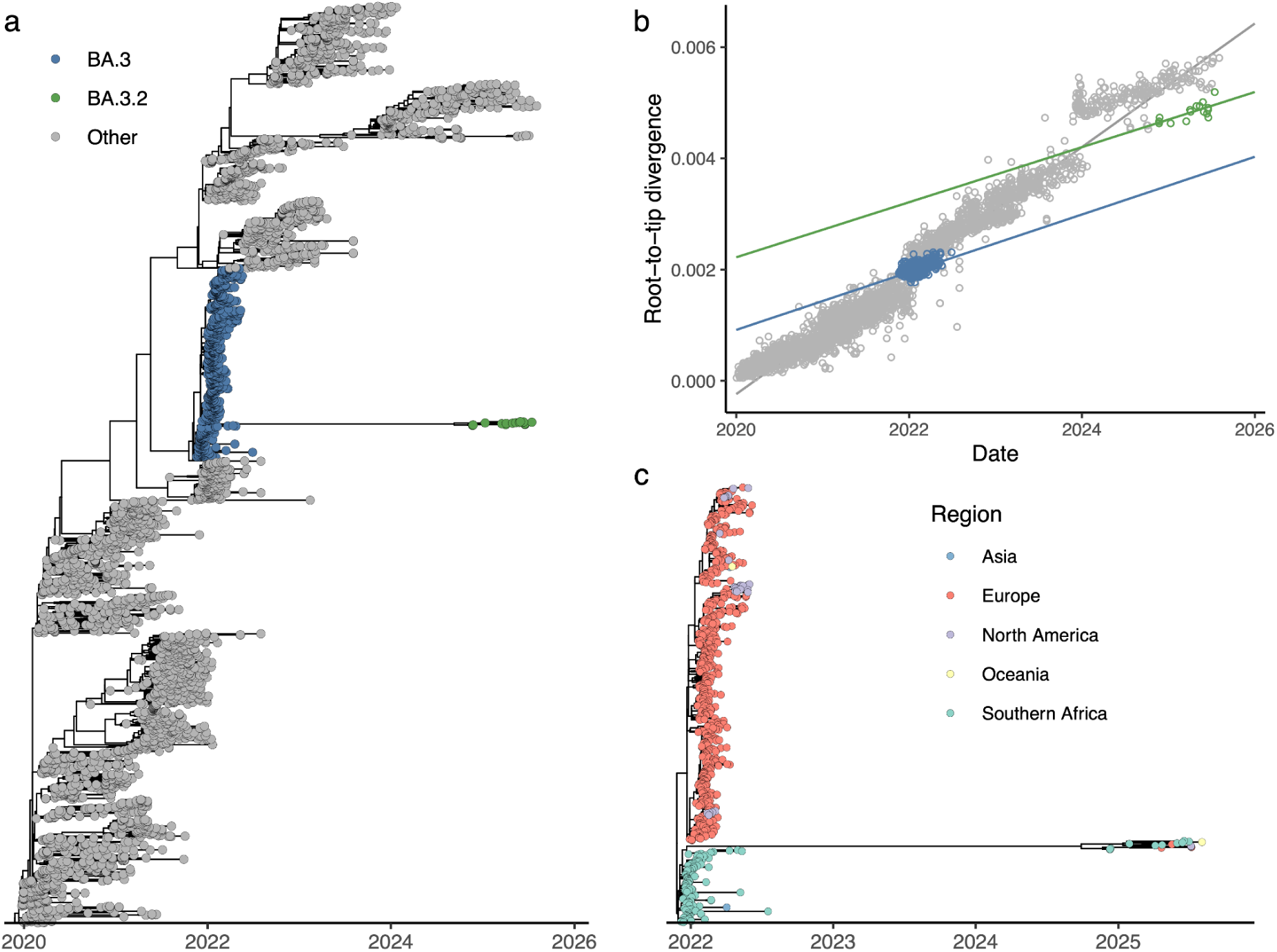
- Phylogenetic analysis of BA.3 lineages in the context of globally subsampled sequences. **a** Time-scaled phylogeny of all BA.3 sequences within a globally representative subset of SARS-CoV-2 sequences (grey), highlighting the long branch separating parental BA.3 sequences (blue) from the more recently emerged BA.3.2 cluster (green). **b** Root-to-tip analysis of high-quality BA.3 sequences within a globally representative subset of SARS-CoV-2 sequences, with ordinary least squares regression lines plotted against the BA.3 parent lineage (blue) and BA.3.2 lineage (green) respectively, highlighting the divergence in projected vs observed number of mutations. **c** Expanded view of the BA.3.2 lineage branching from the BA.3 parent clade, coloured by region, illustrating the phylogenetic relationship between BA.3.2 sequences and the broader BA.3 lineage.

The BA.3.2 sequences exhibit markedly greater genetic divergence than would be expected if BA.3 had continued evolving at the rate predicted from the parent lineage (Figure 2b). This clear disparity between projected and observed divergence confirms the occurrence of a saltation event, characterised by a rapid, punctuated accumulation of mutations rather than the steady, clock-like process typically observed.

Prior to BA.3.2, BA.2 was the only lineage in the VOC era that has given rise to such long-branch, highly divergent saltation-variants with a degree of geographical spread mirroring that seen with BA.3.2. Hundreds of Delta, BA.1, BA.4, and BA.5 sequences with mutation profiles that, in the number and types of spike mutations they possess, resemble widely spreading BA.2 saltation-variants such as BA.2.3.20, BA.2.10.4, BA.2.75, BJ.1 (later XBB after recombination with a BA.2.75 descendant), BA.2.83, BA.2.87.1, BP.1, BS.1, and DD.1, but none spread internationally in the way that these BA.2 lineages did. In this sense BA.3.2 is unique. Given the vastly greater numbers of infections caused by Alpha, Delta, BA.1, BA.4, BA.5, and other VOC descendants, this suggests BA.3.2 may share distinct characteristics with BA.2 that are absent from all other VOC.

To better assess the geographical origin on the BA.3.2 lineage we constructed a phylogeny including all BA.3-related sequences (Figure 2c). This phylogeny shows BA.3.2 branching from a cluster of southern African BA.3 genomes, confirming its regional origin and evolutionary continuity with the earliest Omicron lineages.

### Evolutionary diversification and geographic spread of BA.3.2

Time-calibrated Bayesian phylogenetic analysis of BA.3.2 sequences (Figure 3a) reveals additional phylogenetic structure, with the lineage subdividing into at least two well-supported sublineages corresponding to BA.3.2.1 and BA.3.2.2. In late 2025, BA.3.2.2 became the dominant branch of BA.3.2 globally and composed approximately 30% of all SARS-CoV-2 sequences collected in Germany and Western Australia by late November. Following its inferred emergence into the general human population around May 2024 (95% highest posterior density (HPD), February 2024 to September 2024), BA.3.2 persistently circulated in southern Africa during its divergence into the BA.3.2.1 lineage by late September 2024 (95% HPD August to November 2024) and the BA.3.2.2 lineage by early October 2024 (95% HPD July to December 2024). These timings are consistent with estimates reported by Jule *et al*.^11^, who likewise depicted BA.3.2 as phylogenetically related to a cluster of southern African BA.3 genomes.

**Figure 3.**
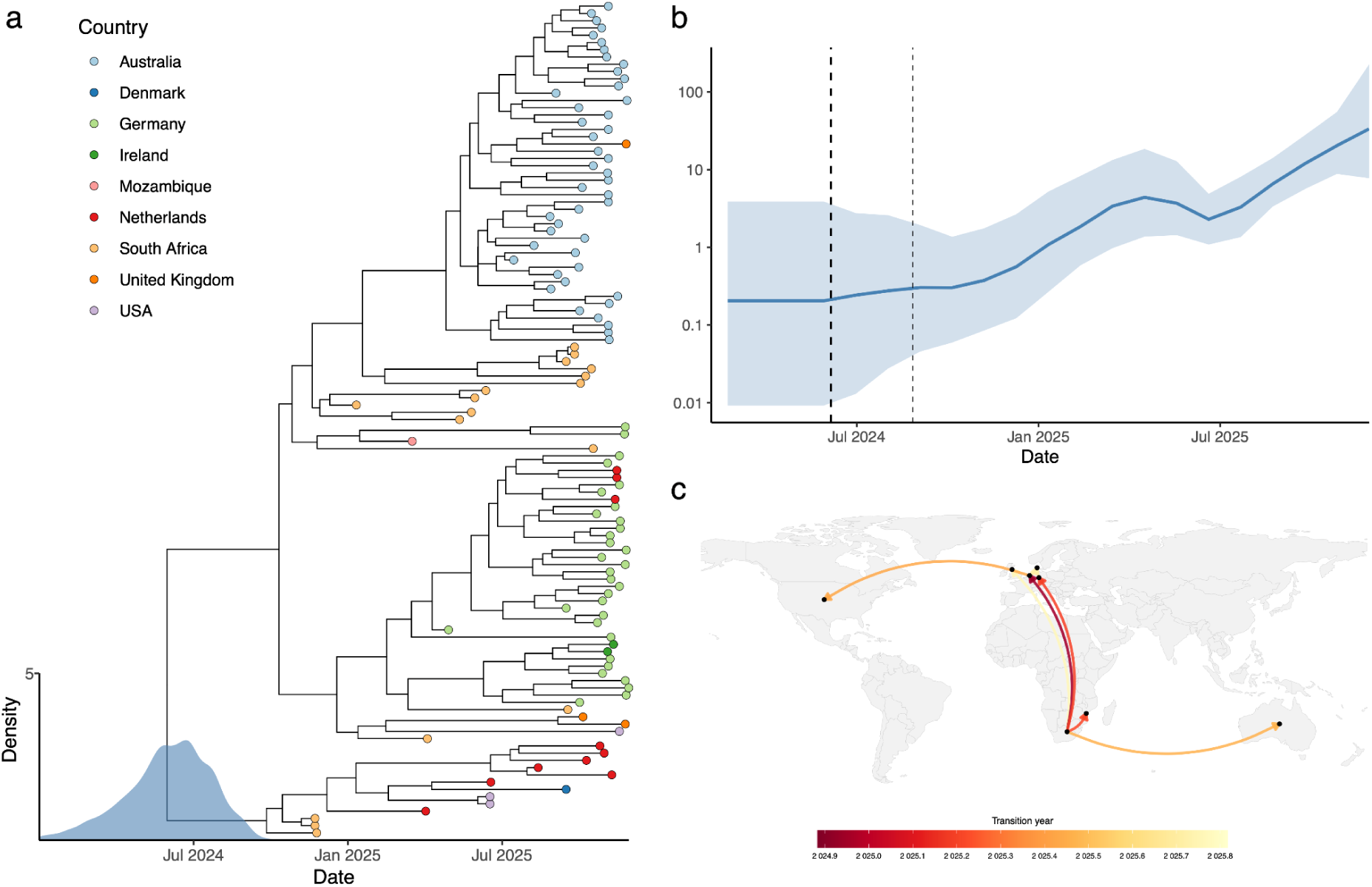
- Phylogenetic, phylodynamic, and phylogeographic characterisation of the BA.3.2 lineage as of December 2025. **a** Time-scaled maximum clade credibility (MCC) tree of BA.3.2 sequences with tip colours indicating sampling country. **b** Bayesian skyline plot showing the effective population size (Ne) through time, highlighting an initial expansion, subsequent decline and regrowth of the lineage from July 2025. **c** Discrete phylogeographic reconstruction of BA.3.2 through November 2025, illustrating inferred country-level movements. Arc colour denotes the relative timing of transitions, with directionality represented anti-clockwise.

The inferred demographic history of BA.3.2 showed an initial increase in effective population size (Ne) following emergence peaking in April 2025, a subsequent decline through to July, and an increase again thereafter (Figure 3b).

Phylogeographic reconstructions position South Africa as the primary source of multiple independent BA.3.2 exportation events during the lineage’s early expansion, with inferred transitions to the Netherlands in late 2024, to Mozambique in early 2025, and subsequently to Germany and Australia around March and June 2025, respectively (Figure 3c). However, these inferred movements likely underrepresent the true timing and scale of transmission and spatial spread. In the six-month period from 1 November 2024 to 30 April 2025, just 398 SARS-CoV-2 sequences were collected from southern African countries, out of a total of 134,401 global GISAID sequences (0.30%). Of these 398 sequences, 366 (92.0%) originated from South Africa, including 10 of the 11 BA.3.2 sequences collected in southern Africa during this period.

A single transition from the Netherlands into the United States was inferred in June 2025, corresponding to an imported case with supporting travel history, with no further detections until November 2025, when BA.3.2 was identified through wastewater surveillance across multiple U.S. states^12^. By December 2025 over half of the available BA.3.2 lineages had been obtained from Australia, where it was first detected on July 15, 2025 (Figure S2c). While sustained circulation has occurred in Western Australia, where detections have been most frequent, the lineage has shown only modest expansion in that region (Figure S2d). Renewed inferred movements across Europe, particularly in September through November 2025, mark continuing global spread, with up to 10% of sampled SARS-CoV-2 genomes collected in Germany during November 2025 being BA.3.2 (Figure S2b).

Although BA.3.2 currently accounts for only a small proportion of globally sampled sequences (2.16% of sequences sampled in November 2025; Figure S1), suggesting more limited growth relative to other circulating variants, its continued detection across multiple regions and evidence of renewed expansion highlight its potential to persist and diversify further, possibly including the acquisition of additional adaptive mutations.

Thirty-nine of BA.3.2’s amino-acid (AA) substitutions are located in the spike protein, along with eleven new AA deletions (Δ136–147 and Δ243–244) and a four-AA insertion (ins214:ASDT) (Figure 4). Perhaps most notably, the BA.3 spike N-terminal domain (NTD) has undergone extensive rearrangement, not unlike BA.2.87.1, another recent saltation-variant originating in South Africa^10,13,14,70^. Four of the spike substitutions involve two non-synonymous nucleotide changes, and the 12-nucleotide insertion contributes to further NTD remodeling.

**Figure 4.**
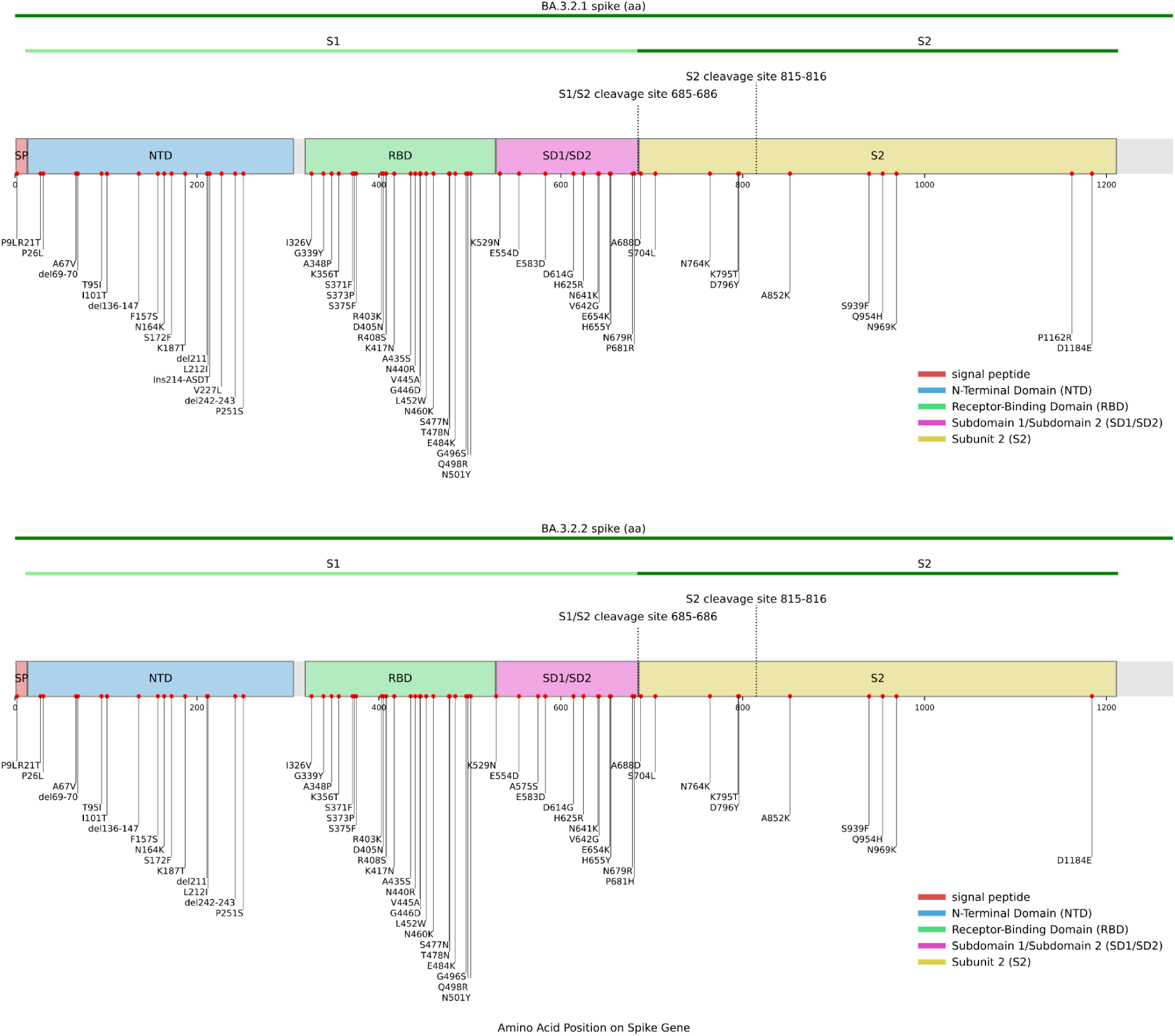
- Mutational profile of BA.3.2.1 and BA.3.2.2 across the spike region highlighting amino acid changes characteristic of each sublineage.

Within the receptor-binding domain (RBD), BA.3.2 carries fifteen amino-acid substitutions, three of which (S446D, L452W, and A484K) arise through two nucleotide changes, a sign of intense selection pressure. Though the original BA.3 lineage had very limited spread (∼760 sequences sampled worldwide) relative to BA.1 and BA.2, it is remarkable that during its short period of circulation, three of the 15 RBD mutations found in BA.3.2 also independently evolved along various different branches of the BA.3 tree, including one 28-sequence branch that acquired G496S and a nine-sequence branch with both D339Y and R408S. BA.3.2 did not descend from either of these branches, suggesting that on a BA.3 background, these mutations confer selective advantages at both an intrahost and interhost level.

### The pattern of substitutions seen at spike sites 405 and 408 is suggestive of epistasis between these sites

Shortly after the emergence of Omicron, an analysis of BA.1 spike mutation patterns identified clusters of mutations that had been strongly selected against during the first two years of the pandemic but which appeared together in BA.1 and which therefore likely interacted epistatically to confer fitness benefits: such as the spike codon 371, 373 and 375 mutations in all Omicron variants^15^. D405N and R408S were both extremely rare mutations in the pre-Omicron era, occurring in just 66 (0.00095%) and 199 (0.0029%) sequences, respectively (see Methods). BA.2, by far the most successful of the three original Omicron variants, had both D405N and R408S, which have been maintained in all subsequent BA.2 lineages. BA.3 had only D405N, and the subsequent evolution of R408S both in ancestral BA.3 and in BA.3.2 also suggests positive epistasis between these two mutations, which is further supported by co-occurrence patterns observed in BA.1 sequences (Supplementary Results S1).

The underlying cause of epistasis between the D405N and R408S mutations is suggested by structural studies of the spike trimer. Cryo-EM analysis of the BA.2 spike shows that R408S eliminates an interprotomer hydrogen bond with F375, which allows F375 to rotate toward the adjacent RBD, possibly improving RBD-RBD packing within the spike trimer. Further, a molecular dynamics study examining the transition of the ancestral Wuhan spike from closed to open conformations, found that in every transition to the “up” state, an interprotomer D405-R408 salt bridge was formed^16^. One might speculate that when either D405 or R408 mutates, this transitional salt bridge is eliminated, and other, compensatory mutations are necessary to establish a viable pathway to the RBD-up state: a state essential for ACE2 binding and infection.

### Reversion mutations likely increase ACE-2 binding and promote escape from neutralizing antibodies

Two of the BA.3.2 RBD mutations are reversions to the Wuhan reference sequence: R493Q and H505Y. In recent antibody escape-mutation analysis, spike residue 505 was identified as the top RBD escape site^17^. R493Q confers a large increase in ACE2 affinity^18^ and was also found in BA.4, BA.5, XAY, XBC, BA.2.10.4, BA.2.75, BA.2.3.20, BS.1, BA.2.77, BA.2.83, BA.2.86, and BA.2.87.1: all, like BA.3.2, highly divergent, anachronistic variants marked by a lack of intermediate sequences. Increased receptor affinity has been shown to effectively evade innate immune responses and T-cell immunity, and therefore seems very likely to be selected for during chronic infections^19^. Furthermore, since a minimal level of ACE2 affinity is required for cell entry, mutations like R493Q enable the acquisition of antibody-evading RBD mutations that reduce ACE2 affinity^20^.

### Deletions and mutations in the NTD of the BA.3.2 spike likely have substantial impacts on the stability and immune evasiveness of spike

Beyond the receptor-binding domain, BA.3.2 also shows extensive remodelling of the spike N-terminal domain (NTD). In addition to the three deletions it inherited from BA.3 (Δ69-70, Δ142-144, and ΔN211), the BA.3.2 spike has eleven more amino acid residues deleted: a 9-AA expansion of the N3-loop deletion (Δ136-147) and a 2-AA deletion in the N5 loop (Δ243-244). Deletions in N3, including the much more modest ΔY144, have previously been shown to evade some classes of antibodies^21^, and in the Beta variant, an N5 deletion similar to that of BA.3.2 was shown to cause a dramatic rearrangement of the N4 loop and even contributed to the evasion of some RBD-directed antibodies^22,23^.

Besides these deletions, the BA.3.2 NTD has eight amino acid substitutions relative to BA.3: P9L, R21T, P26L, I101T, F157S, N164K, S172F, K187T, and P251S.

It is likely that the Spike expressed by BA.3.2 is also missing most of its N1 loop. While the codon sites encoding the N1 loops remain undeleted, the P9L substitution shifts the signal peptide cleavage position from residues 13-14 to residues 21-22^24^, effectively deleting most of the N1 loop. Together with Δ136-147 this erases the C15-C136 NTD disulfide bond. Rearrangement of the NTD due to removal or alteration of the C15-C136 disulfide has been shown to efficiently evade NTD-specific neutralizing antibodies^25–27^. Additional detail on NTD deletions and substitutions is provided in the Supplementary Results S2.

### Mutations clustered in the SD1 and SD2 domains of the BA.3.2 spike likely cause alterations to spike cleavage, fusogenicity and transitions from down to up states

The mechanisms involved in the spike NTD’s modulation of spike cleavage and fusogenicity are not known in detail, but experiments indicate that it involves allosteric interactions among multiple domains and across protomers in the trimeric spike protein^28,29^. The SD2 domain (592-697) is closely associated with the NTD^30–32^, and an interprotomer beta strand consisting of 701-705 and 787-790 has been shown to be a vital link in relaying a fusion-inhibiting signal from the NTD to S2’^28^.

This is intriguing as BA.3.2 possesses, in addition to its extensive NTD modifications, six mutations in the SD2 region (H625R, N641K, V642G, E654K, K679R, A688D), as well as the SD2-adjacent S704L, which is within the aforementioned NTD-S2’-communicating interprotomer beta strand that has a profound effect on membrane fusion activity^28^. Three of these SD2 mutations occur in the structurally crucial 630 loop (619-642), which modulates spike stability and the relative frequencies of the RBD-up and RBD-down conformations, which in turn govern the RBD’s ability to bind to the ACE2 receptor on host cells: an essential step in cell entry^22,33–35^.

A688D has occurred in fewer than 200 SARS-CoV-2 sequences. The introduction of a negatively charged residue in the immediate vicinity of the S1/S2 cleavage site (685/686) seems likely to have major functional effects, perhaps compensating for the instability induced by BA.3.2’s large NTD deletions.

The SD1 domain (529-591) interacts closely with the RBD (334-528), modulating spike stability in both the closed 3-RBD-down and RBD-up states while also relaying signals from the fusion-peptide proximal region (FPPR) and heptad repeat 1 (HR1) regions of S2 to the RBD. The A570D mutation in Alpha, for example, led to new SD1 interactions with K854, resulting in an increase in the frequency of the RBD-up conformation^23,34^. The T572I mutation present in a large number of BA.2.86 descendants, has also been shown, through deep mutational scanning, to increase RBD-up frequency^20^.

BA.3.2 possesses three SD1 mutations: K529N (which likely adds a glycan at N529), E554D, and E583D. Furthermore, it has I326V in the highly conserved NTD-RBD linker (N2R) region (306-334), which is in direct contact with all four S1 regions (NTD, RBD, SD1, SD2)^31,36^. The precise effects of the large number of SD1 and SD2 mutations present in BA.3.2 likely indicate extensive fine-tuning of BA.3.2 spike stability and conformational dynamics.

### Similarities between BA.3.2 and other highly divergent sequences suggest that some of the S2 mutations in BA.3.2 may be adaptive

The five BA.3.2 S2 mutations are S704L, K795T, A852K, S939F, and D1184E. Interestingly, approximately 7% of BA.2.12.1 sequences, one of three other major Pango variants with S704L, also had a N164K mutation. S939F is in BA.2.86 and was not uncommon in circulation prior to 2023. The remaining three BA.3.2 S2 mutations, K795T, A852K, and D1184E, are extremely rare, having occurred in only 51, 223, and 32 sequences, respectively which, on its own, may suggest that collectively these mutations may be epistatically interacting to yield a fitness advantage.

Many spike mutations in the BA.3.2 branch have been seen before in other major lineages. These convergent, and therefore likely adaptive, mutations include the S2 mutations, S704L and S939F (the other convergent spike mutations being R21T, F157S, R403K, R408S, A435S, L452W, N460K, A484K, R493Q, G496S, and K679R). The rarity of K795T, A852K, and D1184E mutations makes it impossible to glean information on their potential fitness costs or benefits based only on examination of available SARS-CoV-2 genome sequence data.

Nevertheless, examination of highly divergent, long-branch sequences that share rare mutations with BA.3.2 reveals a number of striking patterns that undoubtedly contain clues that could help unveil the functional and evolutionary strategies of BA.3.2 and reveal how the different domains of the spike protein interact more generally during cell entry and throughout the viral life cycle.

Among the non-BA.3.2 sequences carrying an A852K mutation, are ten that are found in six distinct, highly divergent lineages (i.e. each of the six lineages has ≥10 private spike amino acid substitutions and A852K). Furthermore, like BA.3.2, each of these sequences is marked by NTD deletions, most of them extensive (see table S1). Notably, all six of these lineages are descendants of BA.1, which has an NTD identical to that of BA.3: an indication of the intimate connection between the spike NTD and S2 regions that are involved in spike cleavage and membrane fusion. Another unique characteristic of BA.1 is the N856K mutation, which has been linked to stabilization of SD1 through the creation of hydrogen bonds with either D568 or T572I^37^ or a new salt bridge with D568^33^, possibly contributing to a more stable three-RBD-down conformation and increased antibody evasion^20,38^. On a BA.1 spike background, A852K creates a stretch of closely spaced lysine residues at 852-854-856. BA.2 lacks N856K but in the BA.2 three-RBD-down spike structure (PDB 7Ub0)^36^, the A852 side chain appears to point directly at D568, suggesting that the A852K mutation may create a similar bond with D568 as N856K did in BA.1; possibly stabilizing the closed spike conformation.

Only two divergent non-BA.3.2 sequences (i.e. sequences carrying ≥10 spike AA substitutions/deletions and ≥ 20 total AA substitutions/deletions) carry the K795T mutation, and strikingly, each also has independently acquired the S2 mutations D843G and I896T. Like D843G, I896T is a rare mutation (446 total sequences, 0.0029%), so the co-occurrence of three such rare S2 mutations is likely meaningful. One of these K795T sequences is a BA.1 descendant that, like BA.3.2, contains an additional large NTD deletion (Δ150-157 in this case). The other divergent K795T sequence, a BA.5 descendant (BA.5.2.20), has even more astonishing similarities to BA.3.2, including two NTD deletions very similar to those of BA.3.2 (Δ134-138 and Δ243-244), C15-C136 disulfide elimination through P9L and deletion of C136, a four-AA insertion in the exact same location as the four-AA insertion in BA.3.2 (ins214:PQAQ), R403K, G496S, and V642G (as well as H681R, found in BA.3.2.1). This suggests that the K795T mutation may be adaptive in the context of the extensive NTD rearrangements seen in BA.3.2.

The available SARS-CoV-2 genome sequence data reveals nothing noteworthy about the adaptive value of the S2 mutation, D1184E.

### Similarities between BA.3.2 and BQ1.1.1 implies that there may presently be an open niche for BA.3.2-like lineages

Perhaps the most remarkable similarities to BA.3.2 are exhibited by an extremely divergent BQ.1.1.1 lineage from Canada from early 2025 that has transmitted at least once (GISAID accession numbers EPI_ISL_19807128, EPI_ISL_19795466, EPI_ISL_19807229, EPI_ISL_19860648, EPI_ISL_19860650, EPI_ISL_19881782). Its similarities to BA.3.2 include C15-C136 disulfide deletion through a P9L mutation and the deletion of C136, deletions in N3 and N5 NTD loops (Δ135-139 and Δ241-247), a substitution at R21 (R21K), A67V (inherited by BA.3 but private to the BQ.1.1.1 lineage), R403K, A435S, V445A, G496S, H505Y (reversion), K529N, E554D, V642G, H681R (in BA.3.2.1), a substitution at A688 (A688T), and, most remarkably, a four-AA insertion at nearly the same location as that found in BA.3.2 (ins212:TQAG). It also has the rare D843G (in only 819 sequences, 0.0052%), which, though not present in BA.3.2, is found in seven of the eight divergent sequences (i.e. sequences with ≥10 private spike substitutions) with either of the K795T or A852K mutations.

BQ.1.1.1 and the eight other divergent lineages carrying K795T and/or A852K mutations mentioned in the previous section, all contain combinations of what appear to be related SD2, FCS-region, and S2 mutations, including V642G (3/9), H681R (6/9), T791I (3/9), K795T (2/9), D843G (7.5/9), A852K (6/9), I896T (2.5/9), and D936H/N/G (5/9). The S2 mutations in this list are extremely rare and, whenever they do occur, are accompanied by two distinct features in the NTD regions of the spikes that they are found in. Perhaps, tellingly, both of these features are found in the BA.3.2 spike. All nine lineages contain private NTD deletions, many of which closely resemble the NTD deletions of BA.3.2. Furthermore, all nine lineages have an NTD insertion in the 212-214 region: seven inherited the BA.1 214:EPE insertion, while the other two (both BA.5* variants) acquired unique four-AA insertions. Novel spike insertions are rare, so the presence of insertions at 212-214 in all nine of these lineages, representing three unique convergent evolutionary events, in combination with convergent mutational patterns resembling those seen in the BA.3.2 SD2, FCS and S2 regions, is almost certainly not coincidental.

The exact adaptive value of this mutational combination awaits experimentation, but judging by its repeated emergence and the sustained, international spread of BA.3.2, it seems to represent what is presently a successful survival strategy for SARS-CoV-2: both for sustained infection within one host, and for interhost transmission.

### The adaptive value of the ORF7 and ORF8 deletions in BA.3.2 remain unclear

Outside of spike, the BA.3.2 genome has changed remarkably little in the three years since BA.3 ceased circulating, with one major exception: ORF7a, ORF7b, and ORF8 are entirely deleted from BA.3.2. The deletion spans 871 nucleotide positions (27,380-28,252)^11^, and closely resembles the ORF7a-ORF7b-ORF8 deletions of GW.5.1, FW.1.1, GE.1.2, and several other “short” XBB variants that circulated in late 2023 before the JN.1 sweep^39^. Inspection of the underlying read-level data indicates that apparent partial or complete reversion of the ORF7–ORF8 region in some genomes is most consistent with alignment or assembly artefacts, rather than true biological restoration of this region.

Mutations that reduce or abolish ORF7 and ORF8 expression have been common throughout the pandemic^40–42^, with some tentative evidence showing that certain specific ORF8 deletions could reduce virulence^43,44^. However, fitness reductions from deletions in ORF8 may be more likely to result from disruption of genomic and subgenomic secondary RNA structure than from loss of the ORF8 protein^45^. The precise nature of the deletion may therefore be crucial in determining its effects on viral fitness and virulence.

### Evolutionary and functional implications of BA.3.2

Together, these findings indicate that BA.3.2 represents a long-divergent descendant of BA.3 that evolved in isolation before re-emerging in late 2024, underscoring the potential for highly divergent SARS-CoV-2 lineages to arise even as global genomic surveillance declines. BA.3.2 carries an extensive set of genomic changes, including 39 amino acid substitutions in the spike glycoprotein, two large N-terminal domain deletions, and a novel four-residue insertion, together with a large deletion spanning ORF7a, ORF7b, and ORF8. The co-occurrence of D405N and R408S, which were both extremely rare prior to Omicron, and re-emerged together in ancestral BA.3 and BA.3.2, suggests epistasis between these sites. Reversion mutations R493Q and H505Y likely increase ACE2 binding affinity and promote escape from neutralising antibodies. The loss of the C15–C136 disulfide bond, together with additional deletions and substitutions across the NTD, likely contribute to extensive structural rearrangements with potential impacts on spike stability and immune evasion. BA.3.2 also exhibits multiple substitutions clustered in the SD1 and SD2 domains, including H625R, N641K, V642G, E654K, K679R, H681R, and A688D, which are predicted to influence spike cleavage, fusogenicity, and transitions between RBD conformational states. In combination, these features distinguish BA.3.2 from all other known Omicron lineages and may underlie its persistence and continued circulation following re-emergence.

While BA.3.2 has already achieved sustained international spread, its long-term trajectory remains uncertain. Previous highly divergent Omicron lineages, such as BA.2.87.1, have shown that extensive genomic mutations do not necessarily result in widespread transmission. However, the current epidemiological context differs substantially from the situation in late 2023, when the global sweep of JN.1 likely constrained the expansion of other divergent lineages. Since then, population immunity has likely continued to broaden through repeated infections with antigenically similar variants, creating an altered immunological landscape that may permit re-emergent variants such as BA.3.2 to persist longer, or even expand, under reduced competition.

As long as BA.3.2 continues to circulate it may acquire transmission-enhancing mutations. BA.3.2 is clearly continuing to explore novel evolutionary paths. For example, in early November 2025 two closely related BA.3.2.2 sequences in Western Australia appeared with half of the FCS-adjacent QTQT repeat (S:675-678) deleted. Deletions at this site have only been seen in ∼0.0001% of previous SARS-CoV-2 sequences, and in BA.3.2.2 the QT deletion occurs within a cluster of six other FCS-adjacent mutations that have never previously been seen together. Whether ongoing changes like these are consequential enough to substantially increase the transmissibility of BA.3.2.2 will only become apparent if, or when, the massively accelerated growth of a new offshoot of the BA.3.2 lineage becomes detectable by the global SARS-CoV-2 genomic surveillance network, such as occurred in late 2023 when the now globally dominant JN.1 lineage emerged following the acquisition of an L455S mutation by BA.2.86.

In conclusion, in spite of decreasing genomic surveillance, BA.3.2 is persevering and growing in absolute numbers and prevalence alongside dominant SARS-CoV-2 lineages. The extensive adaptive mutations and deletions across the spike protein, together with the deletion of the ORF7-ORF8 region, indicates that this lineage retains the potential to become of epidemiological concern.

## Methods

### Clinical specimens

SARS-CoV-2 positive nasal or nasopharyngeal swabs (in viral transport medium) were obtained through syndromic surveillance programs in South Africa for influenza-like illness (ILI) among outpatients and severe respiratory illness (SRI) among hospitalised patients (NICD Weekly respiratory pathogens surveillance report, 2024), as well as specimens from routine diagnostic SARS-CoV-2 PCR testing performed at public and private laboratories in South Africa.

### Whole genome sequencing

RNA was extracted on the Chemagic 360 using a CMG-1049 kit (PerkinElmer, Massachusetts, USA). Sequencing was performed using the Illumina COVIDSeq protocol (Illumina Inc., San Diego, CA, USA), an amplicon-based next-generation sequencing approach, using ARTIC version 5.3.2 SARS-CoV-2 primer pools. Pooled PCR-amplification products were then processed for tagmentation and adapter ligation using IDT for Illumina indexes (Illumina Inc.). Libraries were quantified using a Qubit 4.01 fluorometer (Invitrogen, Waltham, MA, USA) and the Qubit dsDNA High Sensitivity assay according to manufacturer’s instructions. Fragment sizes were analysed using a TapeStation 4200 (Invitrogen). Libraries were pooled and normalised to 1 nM concentration with a 10% PhiX spike-in. Libraries were loaded onto a 300-cycle NextSeq P1/P2 reagent cartridge and run on an Illumina NextSeq 1000/2000 instrument (Illumina Inc.).

### Genome assembly, quality control, lineage and clade calling

Illumina sequencing reads were assembled using the CZID SARS-CoV-2 pipeline (CZID.org), with default parameter settings. Reads were trimmed with Trim Galore (Phred <20, length <20 bp), realigned, primer-trimmed with iVar and aligned to the SARS-CoV-2 reference genome (accession ID: MN908947.3) using minimap2. iVar was also used to generate a consensus genome requiring ≥10× depth per base. Variants were called using SAMtools/BCFtools and quality metrics assessed with QUAST. Nextclade^47^ and Pangolin v4.3.1^49^ were used for clade and lineage assignments respectively. Nextclade was also used for visualisation of the sequences and the identification of mutations.

### Genomic data acquisition

We retrieved all globally available SARS-CoV-2 sequences designated as BA.3 or its descendant lineages (i.e., BA.3.1 and BA.3.2) from the GISAID database (https://gisaid.org) as of 10 December 2025. This included a total of 1389 sequences, as assigned by the Phylogenetic Assignment of Named Global Outbreak Lineages (PANGOLIN) method^46^.

Sequence alignment and quality assessment were performed using Nextstrain’s Nextclade CLI^47,48^. We retained only sequences with complete collection dates, high genome coverage (>90%), and that passed Nextclade’s internal quality-control metrics. BA.3.2 genomes were the only exception: all 135 available BA.3.2 sequences were initially retained to maximise representation for descriptive assessments, but lower-coverage or QC-failing BA.3.2 genomes were subsequently excluded from phylogenetic and phylogeographic analyses (resulting in 115 high-quality BA.3.2 sequences). After filtering, a total of 875 sequences were retained for downstream analyses, comprising 760 BA.3 (including BA.3.1) genomes and 115 BA.3.2 genomes.

To construct a global phylogenetic context for the BA.3 and BA.3.2 genomes, we used Nextstrain’s publicly available global dataset, which represents a curated and temporally representative global subsample of SARS-CoV-2 genomes drawn from GenBank^48^. We obtained the full metadata and extracted all entries with associated GenBank accession numbers. The corresponding genomes were retrieved from NCBI via the Entrez efetch API and combined with our locally hosted BA.3 and BA.3.2 genomes. All sequences were standardised and matched to their associated metadata and processed through the Nextstrain preprocessing workflow, including alignment and QC assessment with Nextclade.

### Phylogenetic and phylogeographic analysis

Maximum-likelihood (ML) phylogenies were inferred for the complete dataset and for each BA.3 sub-lineage using IQ-TREE version 2.3.2^50^. The GTR+F+R model of nucleotide substitution, as identified by ModelFinder^51^, was applied and branch support was assessed with the ultrafast bootstrap approximation (1000 replicates) to ensure statistical robustness^52^.

Temporal molecular clock signal and outlier detection were assessed using root-to-tip regression analysis in TempEst^53^. The best-fitting root was selected to maximise the correlation between genetic divergence and sampling time. One sequence (EPI_ISL_17387518) with an anomalously large residual, likely due to an incorrectly recorded collection date, was identified and removed, after which no further substantial clock violations were observed.

A time-scaled phylogeny of the complete BA.3 dataset (including BA.3.2) was first generated in TreeTime^54^ using sampling dates to calibrate branch lengths and estimate divergence times. This analysis confirmed that enforcing a strict molecular clock across the full BA.3 dataset resulted in violations of clock-like behaviour, indicating that the BA.3 lineage group was not evolving under a single, well-calibrated rate. To investigate this further, all BA.3 and BA.3.2 genomes were embedded within a broader global phylogenetic framework, as described in the data acquisition section, and processed using the Nextstrain SARS-CoV-2 workflow, which performs standardised alignment, quality filtering, maximum-likelihood inference, and time-scaling to generate a calibrated global tree. We then extracted genetic divergence statistics from the global tree and performed a root-to-tip analysis to assess temporal signal. Ordinary least-squares regression lines were fitted separately for the parental BA.3 lineage and extrapolated to estimate their expected divergence at the time of BA.3.2 emergence within the context of the global dataset.

Evolutionary dynamics of BA.3.2 were further investigated using Bayesian phylogenetic inference in BEAST v1.10.4^55^. We applied a GTR+I+G substitution model, as identified by ModelFinder, together with an uncorrelated lognormal relaxed molecular clock and a Skygrid coalescent prior to estimate temporal changes in effective population size. Two independent MCMC chains were run for 50 million steps, sampling every 5,000 steps, with 10% discarded as burn-in. Convergence and adequate sampling (ESS > 200) were verified in Tracer v1.7.2^56^, and maximum clade credibility (MCC) trees were summarised using TreeAnnotator.

Spatial dynamics of BA.3.2 were inferred using a discrete phylogeographic model in BEAST, using the same evolutionary and coalescent priors to maintain consistency. Country-level locations were treated as discrete traits, and transitions between locations were estimated under a symmetric diffusion model with Bayesian stochastic search variable selection (BSSVS) to identify well-supported migration pathways. The resulting phylogeographic MCC tree was used to reconstruct the dispersal history of BA.3.2.

### Mutational analysis

All sequences on GISAID that carried the A852K mutation were examined manually. Low quality sequences (with obvious sequencing errors, such as <40% spike coverage, >1000 private mutations, >400 AA deletions in spike, insertions of >30 nucleotides in S2, frameshift-causing S2 insertions of 28-29 nucleotides in the immediate vicinity of S:852) were excluded, and the remaining sequences were used to create an ndjson file using Nextclade CLI with the Wuhan reference sequence, GISAID accession EPI_ISL_402124. The excluded sequences were EPI_ISL_2544196, EPI_ISL_12291325, EPI_ISL_13390549, EPI_ISL_13390572, EPI_ISL_13390585, EPI_ISL_17372611. The numbers of private amino acid substitutions and private deletions were determined for each sequence represented in the ndjson file (both in Spike alone and in the whole genome) using custom Julia code that can be found at https://github.com/ryhisner/BA3_2__code. The same process was carried out for sequences carrying the K795T mutation, except that two artifactual frameshift errors were manually fixed for sequence EPI_ISL_18908924, the first by deleting two “N”s at nucleotide position 21976 (D138), and the second by deleting nucleotides TGNNG at the N:30-33 deletion site: an extremely common sequencing error that occurs at this site.

The statistics for the total numbers and percentages of SARS-CoV-2 sequences with spike mutations K795T, D843G, I896T, and D1184E were determined using CovSpectrum. Advanced filters requiring a Nextclade QC score of 99 or below and with sequencing coverage ≥ 0.9 were utilized. In each search, the denominator for the percentage calculation was determined by the same search but without the mutation of interest. The search queries used were:

- S:K795T & !Nextcladepangolineage:BA.3.2* & S:I794I & [1-of: S:D796D, S:D796Y, S:D796H] & S:F797F
- S:D843G & [2-of: S:G842G, S:I844I]
- S:I896T & [2-of: S:Q895Q, S:P897P]
- !Nextcladepangolineage:BA.3.2* & S:D1184E

In each case, all sequences with the mutation of interest were viewed with Nextclade Web to ensure that each mutation was likely genuine and that no artifactual mutations were included.

The statistics for the total number and percentage of non-Omicron, non-recombinant SARS-CoV-2 sequences with S:D405N or with S:R408S were also determined using CovSpectrum. Advanced filters requiring a Nextclade QC score of 29 or below and with sequencing coverage ≥ 0.9 were utilized, along with the search queries tabulated in Table S1. In each case the numerator for the percentage calculation was determined by the same search but without the mutation of interest.

In each case all sequences with the mutation of interest were viewed with Nextclade Web to ensure that each mutation was genuine and that no artifacts were included. In the “BA.1* with S:D405N and not R408S” search, 107 sequences from the same lab and uploaded on the same day were excluded as these sequences came from dozens of isolated branches on the BA.1 Usher phylogenetic tree^57^, they were therefore judged artifactual.

## Ethics approval

The genomic surveillance efforts of SARS-CoV-2 in South Africa have been approved by Research Ethics Committees at the University of KwaZulu-Natal (BREC/00001510/2020), the University of the Witwatersrand (M180832), Stellenbosch University (N20/04/008_COVID-19), the University of Cape Town (383/2020), the University of Pretoria (H101/17) and the University of the Free State (UFS-HSD2020/1860/2710). Individual participant consent was not required for genomic surveillance. An ethics waiver was obtained from the Human Research Ethics Committee of the University of the Witwatersrand (number R14/49). The ILI and SRI study protocols were approved by local HREC, including the University of Witwatersrand HREC (ILI: M180832 and SRI: M140824), the University of KwaZulu-Natal Human Biomedical Research Ethics Committee (ILI: BF 080/12 and SRI: M496/14), and the University of Cape Town Faculty of Health Science HREC (ILI: 573/2018 and SRI: 836/2014).

## Data availability

All of the SARS-CoV-2 sequences analysed and presented here are publicly accessible through the GISAID platform (https://www.gisaid.org/), using the GISAID identifier: EPI_SET_251217mf

## Code availability

Manuscript materials, including BEAST XML input files and R scripts, are available in our GitHub repository: https://github.com/graeme-dor/SARSCoV2-Omicron-BA.3.2-SouthAfrica and Julia code: https://github.com/ryhisner/BA3_2__code

## Acknowledgements

We gratefully acknowledge all data contributors, i.e., the authors and their originating laboratories responsible for obtaining the specimens, and their submitting laboratories for generating the genetic sequence and metadata and sharing via the GISAID Initiative, on which this research is based. We also thank James Theiler, Will Fischer and Hyejin Yoon for computational support via the COVID-19 Viral Genome Analysis Pipeline at Los Alamos, and Angie Hinrichs for assistance with Pango Lineage designations.

## Funding

Sequencing and modelling activities at KRISP and CERI are supported in part by grants from the Rockefeller Foundation (HTH 017; T.d.O.), the Abbott Pandemic Defense Coalition (APDC; T.d.O.), the National Institute of Health USA (U01 AI151698; T.d.O.) for the United World Antivirus Research Network (UWARN; T.d.O.), the INFORM Africa project through IHVN (U54 TW012041; T.d.O.), the SAMRC South African mRNA Vaccine Consortium (SAMVAC; T.d.O.), the Health Emergency Preparedness and Response Umbrella Program (HEPR Program), managed by the World Bank Group (TF0B8412; T.d.O.), the GIZ commissioned by the Government of the Federal Republic of Germany, the UK’s Medical Research Foundation (MRF-RG-ICCH-2022-100069; T.d.O., H.T.), the Wellcome Trust for the Global.health project (228186/Z/23/Z; T.d.O., H.T.), and the Novo Nordisk Foundation (NNF24OC0094346; H.T.). Bette Korber was supported by the US National Institute of Allergy and Infectious Diseases, National Institutes of Health, Department of Health and Human Services, under Contract No. 75N93021C00015. Syndromic surveillance and SARS-CoV-2 sequencing was supported by the National Institute for Communicable Diseases of the National Health Laboratory Service and the US Centers for Disease Control and Prevention. This work was supported in part by a Fogarty International Centre Global Infectious Disease research training grant from the National Institutes of Health to the University of Pittsburgh and National Institute for Communicable Diseases (D43TW011255). Sequencing activities for NICD are supported by a conditional grant from the South African National Department of Health as part of the emergency COVID-19 response; a cooperative agreement between the National Institute for Communicable Diseases of the National Health Laboratory Service and the United States Centers for Disease Control and Prevention (FAIN# U01IP001048; NU51IP000930); the South African Medical Research Council (SAMRC, project number 96838); the African Society of Laboratory Medicine (ASLM) and Africa Centers for Disease Control and Prevention through a sub-award from the Bill and Melinda Gates Foundation grant number INV-018978, INV-049272 and INV-050051; the UK Foreign, Commonwealth and Development Office and Wellcome (Grant no 221003/Z/20/Z); This work was partly funded by the SEQAFRICA project which is funded by the UK Department of Health and Social Care’s Fleming Fund using UK aid. NICD sequencing was also supported by The Coronavirus Aid, Relief, and Economic Security Act (CARES ACT) through the Centers for Disease Control and Prevention (CDC) and the COVID International Task Force (ITF) funds through the CDC under the terms of a subcontract with the African Field Epidemiology Network (AFENET) AF-NICD-001/2021.

## Conflicts of interest

Nicole Wolter and Anne von Gottberg have received grant funding from the Bill and Melinda Gates Foundation, the US Centers for Disease Control and Prevention and Sanofi. Federico Gueli is an independent researcher (contractor) for Invivyd Inc.

## Notes

### Competing Interest Statement

The authors have declared no competing interest.

