## Supplementary material for "Identification and genomic characterisation of BA.3.2: a highly divergent BA.3-related SARS-CoV-2 lineage from southern Africa"

### Supplementary Information

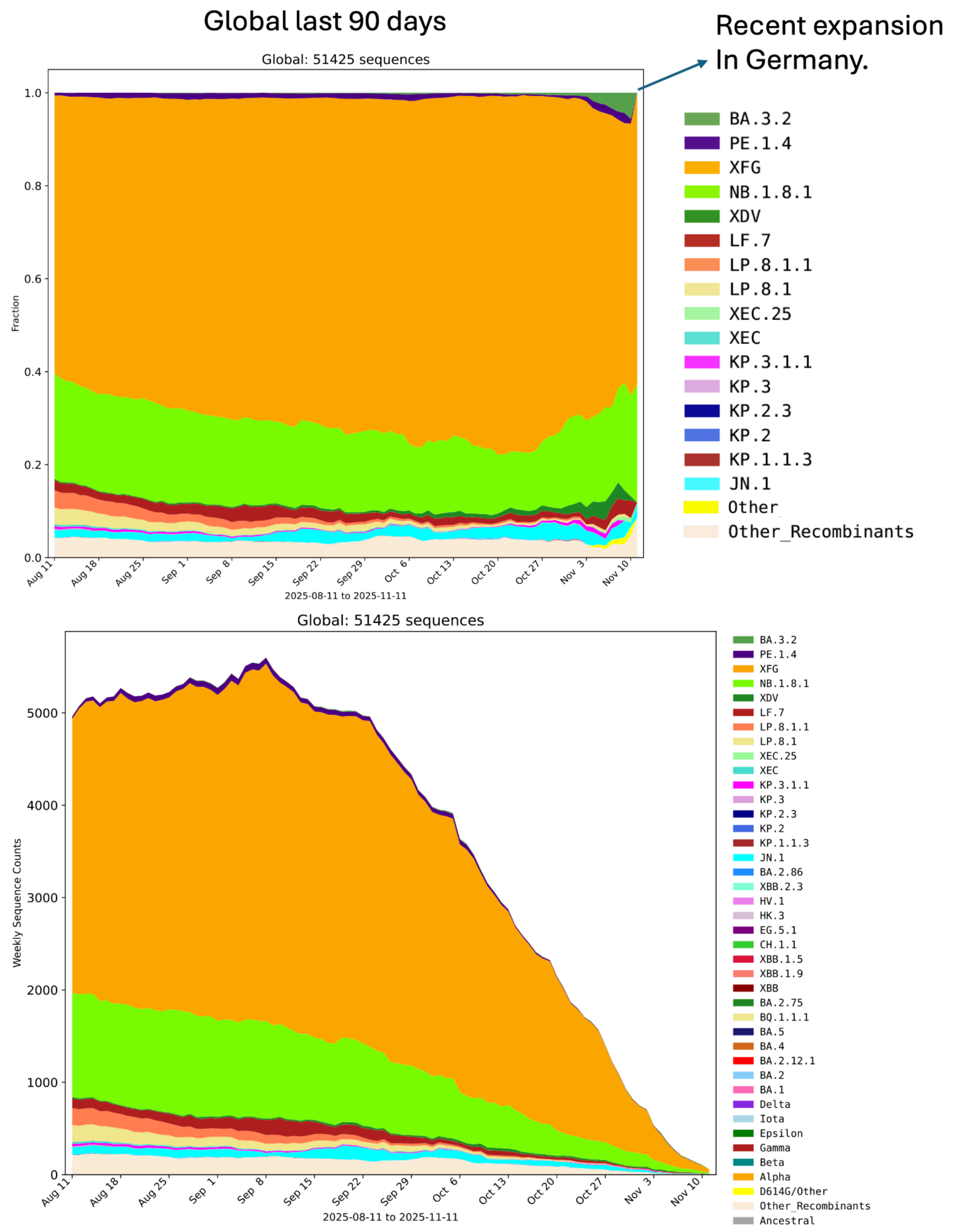


Supplementary figure S1 - Global lineage proportion and volume of SARS-CoV-2 genomes sequenced over the most recent months, highlighting the recent increase in proportion of sequences attributed to BA.3.2

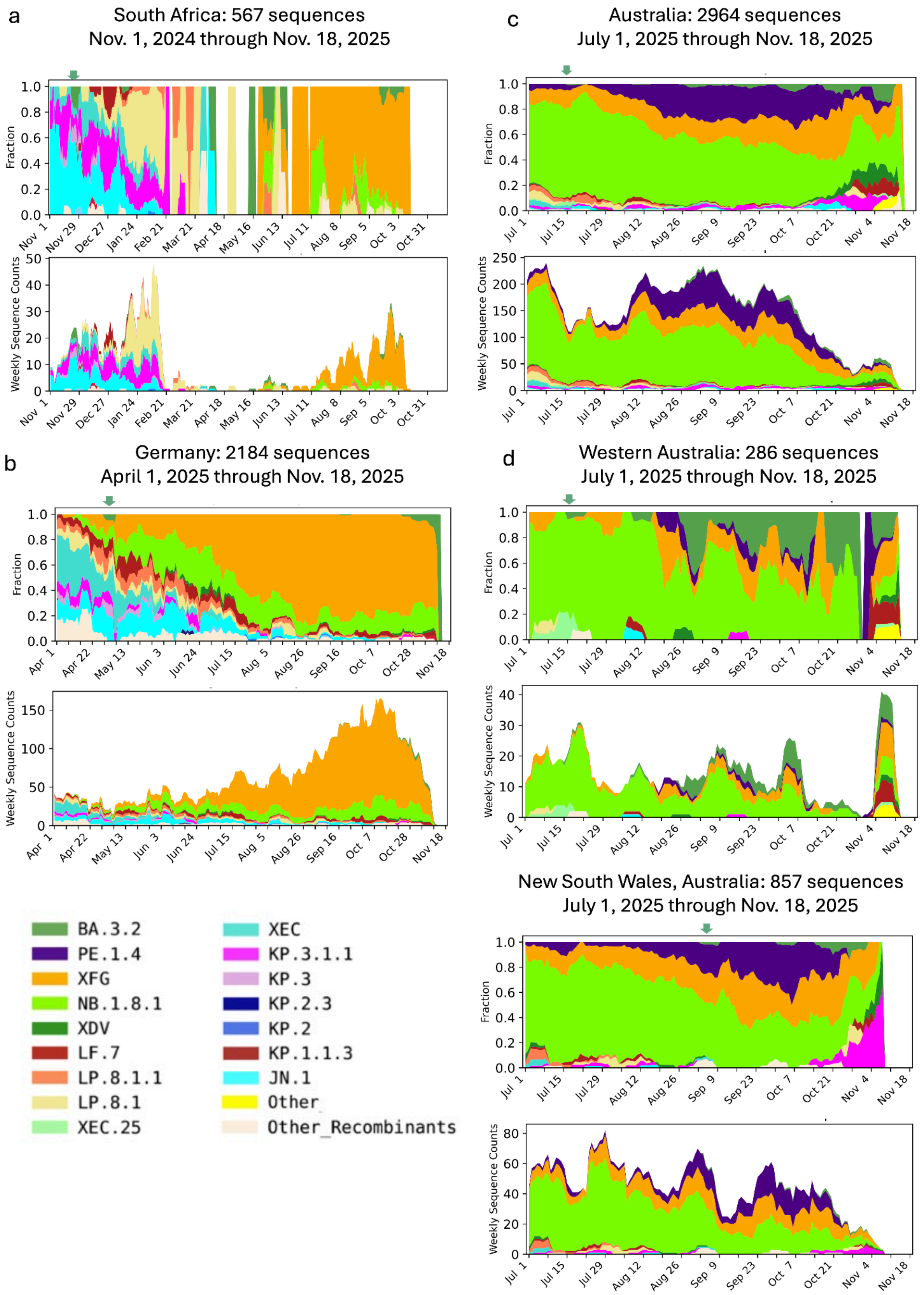
a
South Africa: 567 sequences
Nov. 1, 2024 through Nov. 18, 2025
Fraction
Weekly Sequence Counts
Nov 1 Nov 29 Dec 27 Jan 24 Feb 21 Mar 21 Apr 18 May 16 Jun 13 Jul 11 Aug 8 Sep 5 Oct 3 Oct 31
b
Germany: 2184 sequences
April 1, 2025 through Nov. 18, 2025
Fraction
Weekly Sequence Counts
Apr 1 Apr 22 May 13 Jun 3 Jun 24 Jul 15 Aug 5 Aug 26 Sep 16 Oct 7 Oct 28 Nov 18
c
Australia: 2964 sequences
July 1, 2025 through Nov. 18, 2025
Fraction
Weekly Sequence Counts
Jul 1 Jul 15 Jul 29 Aug 12 Aug 26 Sep 9 Sep 23 Oct 7 Oct 21 Nov 4 Nov 18
d
Western Australia: 286 sequences
July 1, 2025 through Nov. 18, 2025
Fraction
Weekly Sequence Counts
New South Wales, Australia: 857 sequences
July 1, 2025 through Nov. 18, 2025
Fraction
Weekly Sequence Counts
BA.3.2
PE.1.4
XFG
NB.1.8.1
XDV
LF.7
LP.8.1.1
LP.8.1
XEC.25
XEC
KP.3.1.1
KP.3
KP.2.3
KP.2
KP.1.1.3
JN.1
Other
Other_Recombinants

Supplementary figure S2 - **Plots illustrating the frequency of BA.3.2 lineages sampled in the three countries where BA.3.2 has been most commonly detected**. Plots were created using the Geographically Restricted Embers tool at the Los Alamos COVID-19 Viral Genome Pipeline[1] (https://cov.lanl.gov/content/sequence/EMBERS/embers.html) to illustrate the weekly frequency (top) and sequence counts (bottom) of Pango lineage designations over time that were assigned to GISAID[2] data using UShER[3]. Each plot starts with the month BA.3.2 was first detected in a given country and includes all GISAID data available through Nov. 20, 2025 (the last sampling date was Nov. 18 in this set). Pango sub-lineages are grouped into the major lineage designations to enable visualization, for example BA.3.2 is the top lineage in each plot, labeled in green, and includes the BA.3.2.1 and BA.3.2.2 sublineages. The week BA.3.2 was first detected is highlighted with a small green arrow over the "fraction" plots. **a** South African SARS-CoV-2 lineage frequencies. **b** German SARS-CoV-2 lineage frequencies. **c** Australian SARS-CoV-2 lineage frequencies. **d** SARS-CoV-2 lineage frequencies in the two states in Australia where BA.3.2 has been detected as of this sampling.

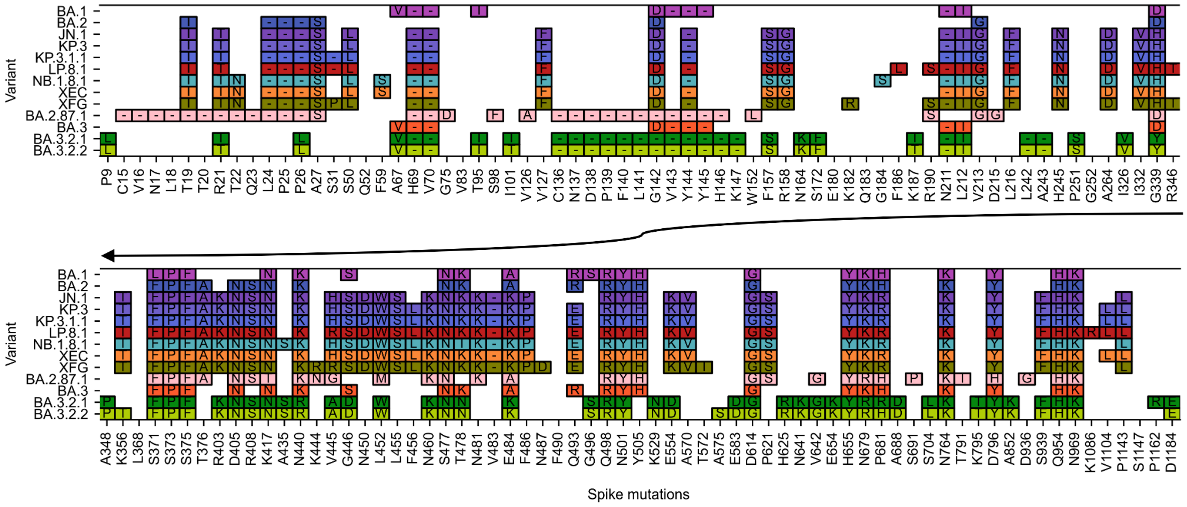


Supplementary figure S3 - Comparative mutation profile of BA.3.2.1 and BA.3.2.2 in relation to other relevant lineages illustrating shared and distinct substitutions.

#### Table S1: Cov-Spectrum Search Queries

| Description | Search |
|---|---|
| non-Omicron, non-recombinant sequences with S:D405N | Advanced Filters — Sequence quality: 0 ≤ Mixed sites score ≤ 101, SNP clusters score ≤ 51, Frame shifts score ≤ 51, Stop codons score ≤ 51, 0.8 ≤ Coverage (from 0 to 1)<br><br>Date Range: 2020-01-06 to 2022-01-01<br><br>S:D405N & [exactly-0-of: ins_S:214:EPE, Nextcladepangolineage:B.1.1.529*, Nextcladepangolineage:XA*, Nextcladepangolineage:XB*, Nextcladepangolineage:XC*, Nextcladepangolineage:XD*, Nextcladepangolineage:XE*, Nextcladepangolineage:XF*, Nextcladepangolineage:XG*, Nextcladepangolineage:XH*, Nextcladepangolineage:XJ*, Nextcladepangolineage:XK*, Nextcladepangolineage:XL*, Nextcladepangolineage:XM*, Nextcladepangolineage:XN*, Nextcladepangolineage:XP*, Nextcladepangolineage:XQ*, Nextcladepangolineage:XR*, Nextcladepangolineage:XS*, Nextcladepangolineage:XT*, Nextcladepangolineage:XU*, Nextcladepangolineage:XV*, Nextcladepangolineage:XW*, Nextcladepangolineage:XY*, Nextcladepangolineage:XZ*, Nextcladepangolineage:XAA*, Nextcladepangolineage:XAB*, Nextcladepangolineage:XAC*, Nextcladepangolineage:XAD*, Nextcladepangolineage:XAE*, Nextcladepangolineage:XAF*, Nextcladepangolineage:XAG*, Nextcladepangolineage:XAH*, Nextcladepangolineage:XAJ*, Nextcladepangolineage:XAK*, Nextcladepangolineage:XAL*, Nextcladepangolineage:XAM*, Nextcladepangolineage:XAN*, Nextcladepangolineage:XAP*, Nextcladepangolineage:XAQ*, Nextcladepangolineage:XAR*, Nextcladepangolineage:XAS*, Nextcladepangolineage:XAT*, Nextcladepangolineage:XAU*, Nextcladepangolineage:XAV*, Nextcladepangolineage:XAW*, Nextcladepangolineage:XAY*, Nextcladepangolineage:XAZ*, Nextcladepangolineage:XBA*, Nextcladepangolineage:XBB*, Nextcladepangolineage:XBC*, Nextcladepangolineage:XBD*, Nextcladepangolineage:XBE*, Nextcladepangolineage:XBF*, Nextcladepangolineage:XBG*, Nextcladepangolineage:XBH*, Nextcladepangolineage:XBJ*, Nextcladepangolineage:XBK*, Nextcladepangolineage:XBL*, Nextcladepangolineage:XBM*, Nextcladepangolineage:XBN*, Nextcladepangolineage:XBP*, Nextcladepangolineage:XBQ*, Nextcladepangolineage:XBR*, Nextcladepangolineage:XBS*, Nextcladepangolineage:XBT*, Nextcladepangolineage:XBU*, Nextcladepangolineage:XBV*, Nextcladepangolineage:XBW*, Nextcladepangolineage:XBY*, Nextcladepangolineage:XBZ*, Nextcladepangolineage:XCA*, Nextcladepangolineage:XCB*, Nextcladepangolineage:XCC*, Nextcladepangolineage:XCD*, Nextcladepangolineage:XCE*, Nextcladepangolineage:XCF*, Nextcladepangolineage:XCG*, Nextcladepangolineage:XCH*, Nextcladepangolineage:XCJ*, Nextcladepangolineage:XCK*, Nextcladepangolineage:XCL*, Nextcladepangolineage:XCM*, Nextcladepangolineage:XCN*, Nextcladepangolineage:XCP*, Nextcladepangolineage:XCQ*, Nextcladepangolineage:XCR*, Nextcladepangolineage:XCS*, Nextcladepangolineage:XCT*, Nextcladepangolineage:XCU*, Nextcladepangolineage:XCV*, Nextcladepangolineage:XCW*, Nextcladepangolineage:XCY*, Nextcladepangolineage:XCZ*, Nextcladepangolineage:XDA*, Nextcladepangolineage:XDB*, Nextcladepangolineage:XDC*, Nextcladepangolineage:XDD*, Nextcladepangolineage:XDE*, Nextcladepangolineage:XDF*, Nextcladepangolineage:XDG*, Nextcladepangolineage:XDH*, Nextcladepangolineage:XDJ*, Nextcladepangolineage:XDK*, Nextcladepangolineage:XDL*, Nextcladepangolineage:XDM*, Nextcladepangolineage:XDN*, Nextcladepangolineage:XDP*, Nextcladepangolineage:XDQ*, Nextcladepangolineage:XDR*, Nextcladepangolineage:XDS*, Nextcladepangolineage:XDT*, Nextcladepangolineage:XDU*, Nextcladepangolineage:XDV*, Nextcladepangolineage:XDW*, Nextcladepangolineage:XDY*, Nextcladepangolineage:XDZ*, Nextcladepangolineage:XEA*, Nextcladepangolineage:XEB*, Nextcladepangolineage:XEC*, Nextcladepangolineage:XED*, Nextcladepangolineage:XEE*, Nextcladepangolineage:XEF*, Nextcladepangolineage:XEG*, Nextcladepangolineage:XEH*, Nextcladepangolineage:XEJ*, Nextcladepangolineage:XEK*, Nextcladepangolineage:XEL*, Nextcladepangolineage:XEM*, Nextcladepangolineage:XEN*, Nextcladepangolineage:XEP*, Nextcladepangolineage:XEQ*, Nextcladepangolineage:XER*, Nextcladepangolineage:XES*, Nextcladepangolineage:XET*, Nextcladepangolineage:XEU*, Nextcladepangolineage:XEV*, Nextcladepangolineage:XEW*, Nextcladepangolineage:XEY*, Nextcladepangolineage:XEZ*, Nextcladepangolineage:XFA*, Nextcladepangolineage:XFB*, Nextcladepangolineage:XFC*, Nextcladepangolineage:XFD*, Nextcladepangolineage:XFE*, Nextcladepangolineage:XFF*, Nextcladepangolineage:XFG*, Nextcladepangolineage:XFH*, Nextcladepangolineage:XFJ*, Nextcladepangolineage:XFK*, Nextcladepangolineage:XFL*, Nextcladepangolineage:XFM*, Nextcladepangolineage:XFN*, Nextcladepangolineage:XFP*, Nextcladepangolineage:XFQ*, Nextcladepangolineage:XFR*, Nextcladepangolineage:XFS*, Nextcladepangolineage:XFT*, Nextcladepangolineage:XFU*, Nextcladepangolineage:XFV*, Nextcladepangolineage:XFW*, Nextcladepangolineage:XFY*, Nextcladepangolineage:XFZ*, Nextcladepangolineage:XGA*, Nextcladepangolineage:XGB*, [5-of: G10447A, A27259C, S:V213G, S:G339D, S:S371F, S:S371L, S:S373P, S:S375F, S:T376A, S:R408S, S:K417N, S:N440K, S:S477N, S:T478K, S:E484A, S:Q493R, S:Q498R, S:N679K, S:N764K, S:D796Y, S:N856K, S:Q954H, S:N969K, S:L981F, M:Q19E, N:S413R, ORF3a:T223I, ORF6:D61L, E:T9I, ORF1a:L3027F, ORF1a:T3090I, ORF1a:P3395H, ORF1b:I1566V, ORF1b:T2163I]] |

| | |
|---|---|
| non-Omicron, non-recombinant sequences with R408S | Advanced Filters — Sequence quality: 0 ≤ Mixed sites score ≤ 101, SNP clusters score ≤ 51, Frame shifts score ≤ 51, Stop codons score ≤ 51, 0.8 ≤ Coverage (from 0 to 1)<br><br>S:R408S & [exactly-0-of: ins_S:214:EPE, C22792T, Nextcladepangolineage:B.1.1.529*, Nextcladepangolineage:XA*, Nextcladepangolineage:XB*, Nextcladepangolineage:XC*, Nextcladepangolineage:XD*, Nextcladepangolineage:XE*, Nextcladepangolineage:XF*, Nextcladepangolineage:XG*, Nextcladepangolineage:XH*, Nextcladepangolineage:XJ*, Nextcladepangolineage:XK*, Nextcladepangolineage:XL*, Nextcladepangolineage:XM*, Nextcladepangolineage:XN*, Nextcladepangolineage:XP*, Nextcladepangolineage:XQ*, Nextcladepangolineage:XR*, Nextcladepangolineage:XS*, Nextcladepangolineage:XT*, Nextcladepangolineage:XU*, Nextcladepangolineage:XV*, Nextcladepangolineage:XW*, Nextcladepangolineage:XY*, Nextcladepangolineage:XZ*, Nextcladepangolineage:XAA*, Nextcladepangolineage:XAB*, Nextcladepangolineage:XAC*, Nextcladepangolineage:XAD*, Nextcladepangolineage:XAE*, Nextcladepangolineage:XAF*, Nextcladepangolineage:XAG*, Nextcladepangolineage:XAH*, Nextcladepangolineage:XAJ*, Nextcladepangolineage:XAK*, Nextcladepangolineage:XAL*, Nextcladepangolineage:XAM*, Nextcladepangolineage:XAN*, Nextcladepangolineage:XAP*, Nextcladepangolineage:XAQ*, Nextcladepangolineage:XAR*, Nextcladepangolineage:XAS*, Nextcladepangolineage:XAT*, Nextcladepangolineage:XAU*, Nextcladepangolineage:XAV*, Nextcladepangolineage:XAW*, Nextcladepangolineage:XAY*, Nextcladepangolineage:XAZ*, Nextcladepangolineage:XBA*, Nextcladepangolineage:XBB*, Nextcladepangolineage:XBC*, Nextcladepangolineage:XBD*, Nextcladepangolineage:XBE*, Nextcladepangolineage:XBF*, Nextcladepangolineage:XBG*, Nextcladepangolineage:XBH*, Nextcladepangolineage:XBJ*, Nextcladepangolineage:XBK*, Nextcladepangolineage:XBL*, Nextcladepangolineage:XBM*, Nextcladepangolineage:XBN*, Nextcladepangolineage:XBP*, Nextcladepangolineage:XBQ*, Nextcladepangolineage:XBR*, Nextcladepangolineage:XBS*, Nextcladepangolineage:XBT*, Nextcladepangolineage:XBU*, Nextcladepangolineage:XBV*, Nextcladepangolineage:XBW*, Nextcladepangolineage:XBY*, Nextcladepangolineage:XBZ*, Nextcladepangolineage:XCA*, Nextcladepangolineage:XCB*, Nextcladepangolineage:XCC*, Nextcladepangolineage:XCD*, Nextcladepangolineage:XCE*, Nextcladepangolineage:XCF*, Nextcladepangolineage:XCG*, Nextcladepangolineage:XCH*, Nextcladepangolineage:XCJ*, Nextcladepangolineage:XCK*, Nextcladepangolineage:XCL*, Nextcladepangolineage:XCM*, Nextcladepangolineage:XCN*, Nextcladepangolineage:XCP*, Nextcladepangolineage:XCQ*, Nextcladepangolineage:XCR*, Nextcladepangolineage:XCS*, Nextcladepangolineage:XCT*, Nextcladepangolineage:XCU*, Nextcladepangolineage:XCV*, Nextcladepangolineage:XCW*, Nextcladepangolineage:XCY*, Nextcladepangolineage:XCZ*, Nextcladepangolineage:XDA*, Nextcladepangolineage:XDB*, Nextcladepangolineage:XDC*, Nextcladepangolineage:XDD*, Nextcladepangolineage:XDE*, Nextcladepangolineage:XDF*, Nextcladepangolineage:XDG*, Nextcladepangolineage:XDH*, Nextcladepangolineage:XDJ*, Nextcladepangolineage:XDK*, Nextcladepangolineage:XDL*, Nextcladepangolineage:XDM*, Nextcladepangolineage:XDN*, Nextcladepangolineage:XDP*, Nextcladepangolineage:XDQ*, Nextcladepangolineage:XDR*, Nextcladepangolineage:XDS*, Nextcladepangolineage:XDT*, Nextcladepangolineage:XDU*, Nextcladepangolineage:XDV*, Nextcladepangolineage:XDW*, Nextcladepangolineage:XDY*, Nextcladepangolineage:XDZ*, Nextcladepangolineage:XEA*, Nextcladepangolineage:XEB*, Nextcladepangolineage:XEC*, Nextcladepangolineage:XED*, Nextcladepangolineage:XEE*, Nextcladepangolineage:XEF*, Nextcladepangolineage:XEG*, Nextcladepangolineage:XEH*, Nextcladepangolineage:XEJ*, Nextcladepangolineage:XEK*, Nextcladepangolineage:XEL*, Nextcladepangolineage:XEM*, Nextcladepangolineage:XEN*, Nextcladepangolineage:XEP*, Nextcladepangolineage:XEQ*, Nextcladepangolineage:XER*, Nextcladepangolineage:XES*, Nextcladepangolineage:XET*, Nextcladepangolineage:XEU*, Nextcladepangolineage:XEV*, Nextcladepangolineage:XEW*, Nextcladepangolineage:XEY*, Nextcladepangolineage:XEZ*, Nextcladepangolineage:XFA*, Nextcladepangolineage:XFB*, Nextcladepangolineage:XFC*, Nextcladepangolineage:XFD*, Nextcladepangolineage:XFE*, Nextcladepangolineage:XFF*, Nextcladepangolineage:XFG*, Nextcladepangolineage:XFH*, Nextcladepangolineage:XFJ*, Nextcladepangolineage:XFK*, Nextcladepangolineage:XFL*, Nextcladepangolineage:XFM*, Nextcladepangolineage:XFN*, Nextcladepangolineage:XFP*, Nextcladepangolineage:XFQ*, Nextcladepangolineage:XFR*, Nextcladepangolineage:XFS*, Nextcladepangolineage:XFT*, Nextcladepangolineage:XFU*, Nextcladepangolineage:XFV*, Nextcladepangolineage:XFW*, Nextcladepangolineage:XFY*, Nextcladepangolineage:XFZ*, Nextcladepangolineage:XGA*, Nextcladepangolineage:XGB*, [5-of: G10447A, A27259C, S:V213G, S:G339D, S:S371F, S:S371L, S:S373P, S:S375F, S:T376A, S:D405N, S:K417N, S:N440K, S:S477N, S:T478K, S:E484A, S:Q493R, S:Q498R, S:N679K, S:N764K, S:D796Y, S:N856K, S:Q954H, S:N969K, S:L981F, M:Q19E, N:S413R, ORF3a:T223I, ORF6:D61L, E:T9I, ORF1a:L3027F, ORF1a:T3090I, ORF1a:P3395H, ORF1b:I1566V, ORF1b:T2163I]] |
| BA.1* with R408S and not S:D405N | Nextcladepangolineage:BA.1* & R408S & [54-of: S:S371L, S:S373P, S:S375F, S:T376T, S:Y380Y, S:G381G, S:V382V, S:S383S, S:K386K, S:L387L, S:N388N, S:D389D, S:L390L, S:C391C, S:F392F, S:T393T, S:N394N, S:V395V, S:Y396Y, S:A397A, S:D398D, S:S399S, S:F400F, S:V401V, S:I402I, S:G404G, S:D405D, S:E406E, S:V407V, S:Q409Q, S:I410I, S:A411A, S:P412P, S:G413G, S:Q414Q, S:T415T, S:G416G, S:K417N, S:I418I, S:A419A, S:Y421Y, S:N422N, S:Y423Y, S:K424K, S:L425L, S:P426P, S:D427D, S:D428D, S:F429F, S:G431G, S:C432C, S:V433V, S:N440K, S:G446S] & [exactly-0-of: T670G, C2790T, G4184A, C4321T, C9344T, A9424G, C9534T, C9866T, C10198T, G10447A, C12880T, C15714T, C17410T, C19955T, A20055G, C21618T, T21633-, A21634-, C21635-, C21636-, C21637-, C21638-, C21639-, T21640-, G21641-, G21987A, T22200G, C26060T, C26858T, G27382C, A27383T, T27384C, A29510C] & [exactly-0-of: T670G, C2790T, G4184A, C4321T, C9344T, A9424G, C9534T, C9866T, C10198T, G10447A, C12880T, C15714T, C17410T, C19955T, A20055G, C21618T, T21633-, A21634-, C21635-, C21636-, C21637-, C21638-, C21639-, T21640-, G21641-, G21987A, T22200G, C26060T, C26858T, G27382C, A27383T, T27384C, A29510C] |

| | |
|---|---|
| BA.1* with S:D405N and not R408S | Nextcladepangolineage:BA.1* & S:D405N & [53-of: S:S371L, S:S373P, S:S375F, S:T376T, S:Y380Y, S:G381G, S:V382V, S:S383S, S:K386K, S:L387L, S:N388N, S:D389D, S:L390L, S:C391C, S:F392F, S:T393T, S:N394N, S:V395V, S:Y396Y, S:A397A, S:D398D, S:S399S, S:F400F, S:V401V, S:I402I, S:G404G, S:E406E, S:V407V, !R408S, S:Q409Q, S:I410I, S:A411A, S:P412P, S:G413G, S:Q414Q, S:T415T, S:G416G, S:K417N, S:I418I, S:A419A, S:Y421Y, S:N422N, S:Y423Y, S:K424K, S:L425L, S:P426P, S:D427D, S:D428D, S:F429F, S:G431G, S:C432C, S:V433V, S:N440K] & [exactly-0-of: T670G, C2790T, G4184A, C4321T, C9344T, A9424G, C9534T, C9866T, C10198T, G10447A, C12880T, C15714T, C17410T, C19955T, A20055G, C21618T, T21633-, A21634-, C21635-, C21636-, C21637-, C21638-, C21639-, T21640-, G21641-, G21987A, T22200G, C26060T, C26858T, G27382C, A27383T, T27384C, A29510C] |
| BA.1* with both S:D405N and R408S | Nextcladepangolineage:BA.1* & S:D405N & [53-of: S:S371L, S:S373P, S:S375F, S:T376T, S:Y380Y, S:G381G, S:V382V, S:S383S, S:K386K, S:L387L, S:N388N, S:D389D, S:L390L, S:C391C, S:F392F, S:T393T, S:N394N, S:V395V, S:Y396Y, S:A397A, S:D398D, S:S399S, S:F400F, S:V401V, S:I402I, S:G404G, S:E406E, S:V407V, R408S, S:Q409Q, S:I410I, S:A411A, S:P412P, S:G413G, S:Q414Q, S:T415T, S:G416G, S:K417N, S:I418I, S:A419A, S:Y421Y, S:N422N, S:Y423Y, S:K424K, S:L425L, S:P426P, S:D427D, S:D428D, S:F429F, S:G431G, S:C432C, S:V433V, S:N440K] & [exactly-0-of: T670G, C2790T, G4184A, C4321T, C9344T, A9424G, C9534T, C9866T, C10198T, G10447A, C12880T, C15714T, C17410T, C19955T, A20055G, C21618T, T21633-, A21634-, C21635-, C21636-, C21637-, C21638-, C21639-, T21640-, G21641-, G21987A, T22200G, C26060T, C26858T, G27382C, A27383T, T27384C, A29510C] & [exactly-0-of: T670G, C2790T, G4184A, C4321T, C9344T, A9424G, C9534T, C9866T, C10198T, G10447A, C12880T, C15714T, C17410T, C19955T, A20055G, C21618T, T21633-, A21634-, C21635-, C21636-, C21637-, C21638-, C21639-, T21640-, G21641-, G21987A, T22200G, C26060T, C26858T, G27382C, A27383T, T27384C, A29510C] |

#### Supplementary Results S1: Evidence for epistasis between spike residues 405 and 408

**This analysis supports the inference of positive epistasis between D405N and R408S described in the main text.**

Here, we quantify the co-occurrence of D405N and R408S in BA.1 sequences to support the inference of epistasis described in the main text. After excluding sequences with any other BA.2 mutations (indicating contamination, coinfection, or recombination) or dropout in the spike region immediately surrounding 405, only five high-quality BA.1 sequences possessed the D405N mutation without R408S (0.0004% of all qualifying BA.1 sequences). A similar search for BA.1 sequences with R408S but without D405N returns 10 sequences (0.0008%). The number of high-quality BA.1 sequences with both D405N and R408S (and no other BA.2 mutations), on the other hand, is 20.

#### Supplementary Results S2: Expanded section on deletions and mutations in the NTD

**Deletions and mutations in the NTD of the BA.3.2 spike likely have substantial impacts on the stability and immune evasiveness of spike (expanded)**

The SARS-CoV-2 NTD is marked by five projecting loops, N1 (14-26), N2 (67-81), N3 (140-158), N4 (173-190), and N5 (241-263)[4]. These loops are exceptionally long in the SARS-CoV-2 Wuhan reference genome compared to other sarbecoviruses[4,5]. In SARS-CoV-2 pseudoviruses, shorter NTD loops have been shown to increase infectivity and fusion activity, but at the expense of greater spike instability and premature S1 shedding, which precludes cell entry[4-6]. The longer NTD loops in SARS-CoV-2 likely enabled, or else were adaptations to, the furin cleavage site (FCS) insertion at the S1/S2 boundary, which is unique among known sarbecoviruses and which makes premature S1 shedding more likely.

In addition to the three deletions it inherited from BA.3 (∆69-70, ∆142-144, and ∆N211), the BA.3.2 spike has eleven more amino acid residues deleted: a 9-AA expansion of the N3-loop deletion (∆136-147) and a 2-AA deletion in the N5 loop (∆243-244). Deletions in N3, including the much more modest ∆Y144, have previously been shown to evade some classes of antibodies[7], and in the Beta variant, an N5 deletion similar to that of BA.3.2 was shown to cause a dramatic rearrangement of the N4 loop and even contributed to the evasion of some RBD-directed antibodies[8,9].

Besides these deletions, the BA.3.2 NTD has eight amino acid substitutions relative to BA.3: P9L, R21T, P26L, I101T, F157S, N164K, S172F, K187T, and P251S.

It is likely that the Spike expressed by BA.3.2 is also missing most of its N1 loop. While the codon sites encoding the N1 loops remain undeleted, the P9L substitution shifts the signal peptide cleavage position from residues 13-14 to residues 21-22[10], effectively deleting most of the N1 loop. Together with ∆136-147 this erases the C15-C136 NTD disulfide bond. Rearrangement of the NTD due to removal or alteration of the C15-C136 disulfide has been shown to efficiently evade NTD-specific neutralizing antibodies[11-13]. Only three lineages with a deleted C15-C136 disulfide, all relatively small, have ever circulated: B.1.640 (~1200 sequences), C.1.2 (~420 sequences), and BA.2.87.1 (~10 sequences).

Similarly rare are the S172F and K187T substitutions, each of which has appeared in only ~400 of the more than 15 million SARS-CoV-2 sequences. K187T likely creates a glycan at N185 and was in the CH.1.1.18 lineage. Over half of the approximately 400 SARS-CoV-2 sequences with S172F were Delta lineages, while fewer than 100 have been found so far in Omicron lineages other than BA.3.2. Interestingly, one highly divergent HP.1.1 (an XBB.1.5 descendant) sequence (EPI_ISL_19808706) with S172F shares several additional spike elements with BA.3.2, including NTD disulfide alteration (through C15Y and W152C), addition of an N99 glycan (through an I101T mutation), ∆243-244, mutation away from S446 (S446N for the HP.1.1, S446D for BA.3.2) and the mutations E583D, V642G, and K679R. Furthermore, like BA.3.2 (which has S:A688D), it has a mutation adjacent to the FCS (T676N, which creates a glycan) that almost certainly inhibits S1/S2 cleavage. The occurrence of such a large number of otherwise uncommon mutations in two unrelated sequences may be an indication that these mutations may be interacting with one another (i.e. via positive epistasis) to yield a fitness advantage.

The addition of three new glycans on the BA.3.2 spike (promoted by I101T, K187T, and K529N), and a fourth glycan in BA.3.2.2, which adds a glycan at N354 via its K356T mutation, could represent a long-term trend in SARS-CoV-2 evolution. The BA.2.86 spike also possessed additional S1 glycans at N245 (via H245N) and N354 (via K356T) relative to previously dominant variants. Furthermore, within a year after its first appearance, a third glycan (either at N22 via T22N or N30 via ∆S31) had become universal among BA.2.86 descendants[14,15], and in spring 2025 over half of all sequences had added another glycan with the R190S mutation. As population immunity grows through repeated vaccination and infection, and antibody breadth continually increases through the affinity maturation process[16,17], the addition of new S1 glycans may represent an increasingly essential immune-evasion strategy for the virus.

#### Data availability

All of the SARS-CoV-2 sequences analysed and presented in the supplementary are publicly accessible through the GISAID platform (https://www.gisaid.org/), using the GISAID identifier: EPI_SET_251123so (Figure S2a); EPI_SET_251123nh (Figure S2b); EPI_SET_251123ve (Figure S2c and S2d).
